# Analyzing Fractal Dimension in Electroconvulsive Therapy: Unraveling Complexity in Structural and Functional Neuroimaging

**DOI:** 10.1101/2024.02.23.24303023

**Authors:** Niklaus Denier, Matthias Grieder, Kay Jann, Sigrid Breit, Nicolas Mertse, Sebastian Walther, Leila M. Soravia, Agnes Meyer, Andrea Federspiel, Roland Wiest, Tobias Bracht

**Affiliations:** Translational Research Center, University Hospital of Psychiatry and Psychotherapy, University of Bern, Bern, Switzerland; Translational Imaging Center (TIC), Swiss Institute for Translational and Entrepreneurial Medicine, Bern, Switzerland; USC Stevens Neuroimaging and Informatics Institute, Keck School of Medicine of USC, University of Southern California, Los Angeles, CA, United States; Institute of Diagnostic and Interventional Neuroradiology, University of Bern, Bern, Switzerland

**Keywords:** electroconvulsive therapy, depression, neuroimaging, functional MRI, neuroplasticity, connectivity

## Abstract

**Background:** Numerous studies show that electroconvulsive therapy (ECT) induces hippocampal neuroplasticity, but findings are inconsistent regarding its clinical relevance. This study aims to investigate ECT-induced plasticity of anterior and posterior hippocampi using mathematical complexity measures in neuroimaging, namely Higuchi’s fractal dimension (HFD) for fMRI time series and the fractal dimension of cortical morphology (FD-CM). Furthermore, we explore the potential of these complexity measures to predict ECT treatment response.

**Methods:** Twenty patients with a current depressive episode (16 with major depressive disorder and 4 with bipolar disorder) underwent MRI-scans before and after an ECT-series. Twenty healthy controls matched for age and sex were also scanned twice for comparison purposes. Resting-state fMRI data were processed, and HFD was computed for anterior and posterior hippocampi. Group-by-time effects for HFD in anterior and posterior hippocampi were calculated and correlations between HFD changes and improvement in depression severity were examined. For baseline FD-CM analyses, we preprocessed structural MRI with CAT12’s surface-based methods. We explored the predictive value of baseline HFD and FD-CM for treatment outcome.

**Results:** Patients exhibited a significant increase in bilateral hippocampal HFD from baseline to follow-up scans. Right anterior hippocampal HFD increase was associated with reductions in depression severity. After applying a whole-brain regression analysis, we found that baseline FD-CM in the left temporal pole predicted reduction of overall depression severity after ECT. Baseline hippocampal HFD did not predict treatment outcome.

**Conclusion:** This pioneering study suggests that HFD and FD-CM are promising imaging markers to investigate ECT-induced neuroplasticity associated with treatment response.

## Introduction

Depression is a highly prevalent mental disorder and globally the leading cause of disability (WHO, 2021). One-third of patients with depression remain resistant to treatment regimens with pharmacotherapy (Rush et al., 2006). More than 50% of these patients respond to treatment with electroconvulsive therapy (ECT) (Group, 2003; van Diermen et al., 2018). ECT-induced volume increase in limbic structures, including bilateral hippocampi and amygdalae, are well-documented findings observed in multiple studies (Bracht et al., 2023; Gryglewski et al., 2021; Nordanskog et al., 2010; Takamiya et al., 2018). Whilst some studies found associations between overall volume increase and clinical improvements (Joshi et al., 2016; Nordanskog et al., 2010), findings of a meta-analysis and a large mega-analysis, suggest that overall volume increase in the hippocampi is unrelated to treatment response (Gryglewski et al., 2021; Oltedal et al., 2018). Consequently, it was proposed that overall enlargements of the entire hippocampus might rather be a non-specific effect of ECT than the primary driver of treatment response. However, other findings suggest that clinical response may be related to structural and functional alterations in specific segments or subcompartments of the hippocampi (Bracht et al., 2023; Leaver et al., 2019; Leaver et al., 2021; Nuninga et al., 2020). Accordingly, it was hypothesized that changes in functional networks of these subcompartments distinguish ECT-responders from non-responders (Leaver et al., 2021). Furthermore, ECT-induced structural changes have not only been identified in the hippocampi but also in hippocampal connection pathways of extended networks related to anterior or posterior hippocampi (Kubicki et al., 2019), which were associated with a disruption of the hippocampus-default mode network (Denier et al., 2023; Gbyl et al., 2024). Thus, for further advancement, novel methods are required that complement and exceed the interpretation of measures that simply assess overall grey matter volume or structural and functional connectivity.

The incorporation of mathematical complexity analyses offer a transformative perspective on existing data, serving as a valuable complementary measure to conventional neuroimaging metrics. Fractal dimension (FD), originally introduced by the renowned mathematician Benoit Mandelbrot in 1967, represents a pivotal concept. It furnishes a quantitative measure of intricacy and self-similarity, opening new avenues for understanding complex neurological phenomena (Mandelbrot, 1967). Fractals are captivating geometric constructs known for their intricate detail and repeated patterns across various scales, maintaining a similar structure regardless of magnification levels (Kenneth, 1990; Mandelbrot and Mandelbrot, 1982). In contrast to traditional Euclidean geometry, which deals with shapes of integer dimensions (e.g., lines [1D], planes [2D], solids [3D]), Fractal Dimension (FD) expands this concept to embrace non-integer dimensions (e.g., dimension of 1. 3̅). This unique property makes FD also a valuable tool for quantifying the complexity and irregularity present in neuroscience measurements (Burns and Rajan, 2015; King et al., 2009). It allows to decipher and model intricate data, finding widespread utility in the analysis of cortical morphology within a clinical context (Meregalli et al., 2022), and also in studies involving patients with depression (Schmitgen et al., 2020; Schmitt et al., 2022). Traditionally, the assessment of FD in cortical morphology (FD-CM) relies on the box-counting method (Madan and Kensinger, 2016). However, more expedient approximation techniques exist, such as the reconstruction of spherical harmonics (Yotter et al., 2011b). Regional FD-CM represents a comprehensive gauge of gyrification, amalgamating data from folding frequency, sulcal depth, convolution of gyral shape, and cortical thickness into a singular metric. This composite measure has the potential to offer insights into dendrite complexity and synaptic density (Im et al., 2006), enriching our understanding of cortical intricacies.

Paralleling efforts in quantifying spatial fractals, Higuchi’s Fractal Dimension (HFD) emerges as a powerful technique for quantifying the intricacy and self-similarity within one-dimensional data, such as time series (Higuchi, 1988; Liehr and Massopust, 2020). In contrast to other nonlinear methods it is considered as highly accurate in estimating FD (Kesić and Spasić, 2016). HFD analysis has proven invaluable in unraveling the complexities and altered connectivity patterns within the brains of individuals affected by various neurological disorders including migraine and neurodegenerative diseases (Djuričić et al., 2023; Garehdaghi and Sarbaz, 2023; Porcaro et al., 2020; Porcaro et al., 2022; Varley et al., 2020). Surprisingly, while HFD has gained traction in exploring depressive disorders, primarily using ECG (George et al., 2023) and EEG data (Kaushik et al., 2023; Kawe et al., 2019; Lord and Allen, 2023), it has remained largely unexplored in the realm of neuropsychiatric disorders and functional MRI data. Transferring HFD to functional neuroimaging presents an intriguing opportunity to harness HFD’s potential for gaining fresh insights into the landscape of these complex conditions.

This is the first study that exploits HFD and FD-CM to explore ECT-induced neuroplasticity through the lens of complexity measurements. We applied these measures in a sample of 20 patients with depression who were scanned before and after an ECT-index series and in 20 healthy controls who were also scanned twice. It was the primary objective of this study to investigate whether ECT induces changes of functional MRI signal complexity in the anterior and posterior hippocampi as assessed with HFD. Secondary outcome measures were associations between changes in HFD and improvements in depression severity and the predictive value of baseline measures of FD-CM and HFD for treatment outcome. In essence, our study sought to introduce FD metrics including FD-CM and HFD into the analysis of depression and ECT treatment, shedding new light on the phenomena of ECT induced neuroplasticity.

## Methods

### Participants

This same sample was used in previous analyses (Bracht et al., 2023; Denier et al., 2023). Baseline measurements of patients with major depressive disorder (MDD) and healthy controls (HC) were also included in larger samples investigating cross-sectional group differences (Bracht et al., 2022a; Bracht et al., 2022c; Denier et al., 2024; Mertse et al., 2022). We recruited 20 patients with a current depressive episode who were scheduled for an ECT-series at the University Hospital of Psychiatry and Psychotherapy Bern. Inclusion criteria were a diagnosis of major depressive disorder (MDD) or bipolar disorder (BD) according to the Diagnostic and Statistical Manual of Mental Disorders (DSM-5) by the American Psychiatric Association (APA, 2013) and age between 18-65 years. Patients with neurological disorders, substance use disorders, psychotic disorders, personality disorders, known claustrophobia, or other contraindications to undergo an MRI scan were excluded. We conducted diagnostic screening using the Mini International Neuropsychiatric Interview (MINI) (Sheehan et al., 1998) and the Structured Clinical Interview for DSM-IV Axis II (SCID-II) (Wittchen et al., 1997). The Edinburgh Handedness Inventory (Oldfield, 1971) was used to assess handedness. Depression severity was assessed using the 21-item Hamilton Rating Scale for Depression (HAMD) (Hamilton, 1967). The depression rating scales were administered both before and after the ECT-index series on the day of the MRI scan. We also included 20 healthy controls (HC) who were matched with the patients regarding age and sex. HC underwent the same assessments. All subjects provided written informed consent, and the study was approved by the local cantonal ethics committee (KEK-number: 2017-00731). For more details of the sample, see table 1.

**Table 1:** Clinical characteristics of groups. See Denier et al., 2023 (Denier et al., 2023).

|  | **ECT**<br>**(n=20)** | **HC**<br>**(n=20)** | **Analyzes** |
| --- | --- | --- | --- |
| **Age (years)** | 44.9 ± 12 | 43.6 ± 14 | p = 0.75 |
| **Sex (female, male)** | 8, 12 | 8, 12 | p = 1.00 |
| **Handedness**<br>**(right, left, ambidextrous)** | 15, 2, 3 | 17, 2, 1 | p = 0.57 |
| **HAMD-21 (Baseline)** | 21.4 ± 5.3 | 0.65 ± 1.0 | p < 0.001 ** |
| **Duration of episode (months)** | 19.8 ± 17 | n/a | n/a |
| **Number of episodes** | 5.2 ± 4 | n/a | n/a |
| **Time between scans (days)** | 52.6 ± 24 | 61.2 ± 17 | p = 0.20 |
| **HAMD-21 (Follow up)** | 10.9 ± 8.1 | 0.25 ± 0.8 | p < 0.001 ** |
| **n (remitter<sup>1</sup>)** | 9 | n/a | n/a |
| **n (responder or remitter<sup>2</sup>)** | 11 | n/a | n/a |
| **n (non-responder)** | 9 | n/a | n/a |

Demographic and clinical variables were compared between ECT and healthy controls using an independent t-tests and $\chi^2$ tests. Abbreviations: ECT: electroconvulsive therapy; HC: healthy controls; 1 remitter: HAMD-21 < 8 points; 2 responder or remitter: HAMD-21 reduction > 50%; n/a: not applicable.

### ECT treatment

The ECT-treatments, using a Thymatron IV system, were conducted in the anesthetic recovery room of the University Hospital in Bern, Switzerland. Most patients (n=17) received right unilateral stimulation as their primary treatment approach. However, among these 17 patients, five switched to bitemporal stimulation, and one patient switched to bifrontal stimulation during the ECT-index series. Additionally, two patients underwent an ECT-index series with bitemporal stimulation, while one patient received bifrontal stimulation. The decisions regarding the initial placement of electrodes and any subsequent switches during the ECT-index series were based on each patient’s clinical presentation and progress. The titration-based method was employed to determine the initial seizure threshold and stimulus intensity. General anesthesia was administered using etomidate, and succinylcholine was utilized for muscle relaxation. The quality of the seizures was monitored using electroencephalogram (EEG) and electromyography (EMG) recordings. The ECT patients received an average of 12.7 ± 4.0 ECT sessions between the MRI scans.

### MRI acquisition

Each participant underwent two MRI scans using a 3 Tesla MRI scanner (Magnetom Prisma, Siemens, Erlangen, Germany) with a 64-channel head and neck coil at the Swiss Institute for Translational and Entrepreneurial Medicine (SITEM) associated with the University Hospital of Bern. The ECT group was scanned before and after an ECT-index series, while the HC group was also scanned at two timepoints, with a similar duration between scans. For the acquisition of T1-weighted data, a bias-field corrected MP2RAGE sequence was employed. The following parameters were used for the MP2RAGE acquisition: field of view (FOV) = 256×256 mm^2^, matrix = 256×256, slices = 256, voxel resolution = 1×1×1 mm³, repetition time (TR)/echo time (TE) = 5000/2.98 ms, inversion time (TI) = 700 ms, and echo time 2 (T2) = 2500 ms. Resting-state functional MRI (rs-fMRI) was acquired using echo planar imaging (EPI) continuously for 8 minutes, with participants in an ‘eyes closed’ condition. The following parameters were used for the acquisition: 480 volumes with 48 slices per volume, FOV = 230×230 mm^2^, matrix = 94×94, voxel resolution = 2.4×2.4×2.4 mm³ isotropic, TR = 1000 ms, and TE = 30 ms.

### Pre-processing of resting-state fMRI

Rs-fMRI pre-processing was conducted using the CONN 21a toolbox (Whitfield-Gabrieli and Nieto-Castanon, 2012) and involved several procedures. First, the EPI volumes were realigned and co-registered to the MP2RAGE images. Segmentation and normalization to the MNI space were then performed, followed by smoothing with an FWHM kernel of 8 × 8 × 8 mm. In contrast to traditional analysis of low frequency fluctuations (0.01 – 0.1 Hz) no band-pass filter was applied to analyze the whole spectrum.

### Calculation of Higuchi’s fractal dimension (HFD) in resting-state fMRI

Higuchi introduced an algorithm to determine the FD of a time series by quantifying the intricacy of waveforms (Higuchi, 1988). We used Matlab R2023a (Mathworks, Natick, Massachusetts) and the higuchi_fractal_dimension.m script of the Complexity toolbox (https://github.com/kayjann/complexity/tree/master) and performed calculations on UBELIX (http://www.id.unibe.ch/hpc), the HPC cluster at the University of Bern.

The signal in each voxel *x* of rs-fMRI with *N* = 480 time points is defined as a time sequence *x*(1), *x*(2), …, *x*(*N*). From this time sequence, we calculated self-similar time sequences 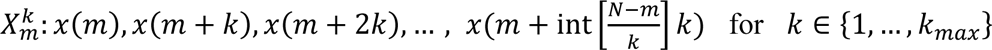 and *m* ∈ {1, …, *k*}. Parameter *k* is the time interval and *k*_*max*_ is a free parameter which we defined as *k_max_* = 5. The formula 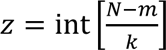 is defined as the upper border of total in time intervals of a sequence with length *k* with int as the integer part of the fraction. For each time sequence 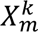 we computed the length 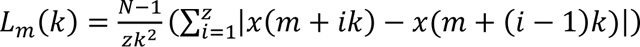. The mean curve length for each time interval *k* = 1, …, *k*_*max*_ was calculated as 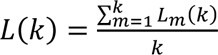. HFD is defined as the best fitting function the double logarithmic dataset {ln(*L*(*k*)), ln (1/*k*)}. See figure 1.

**Figure 1:**
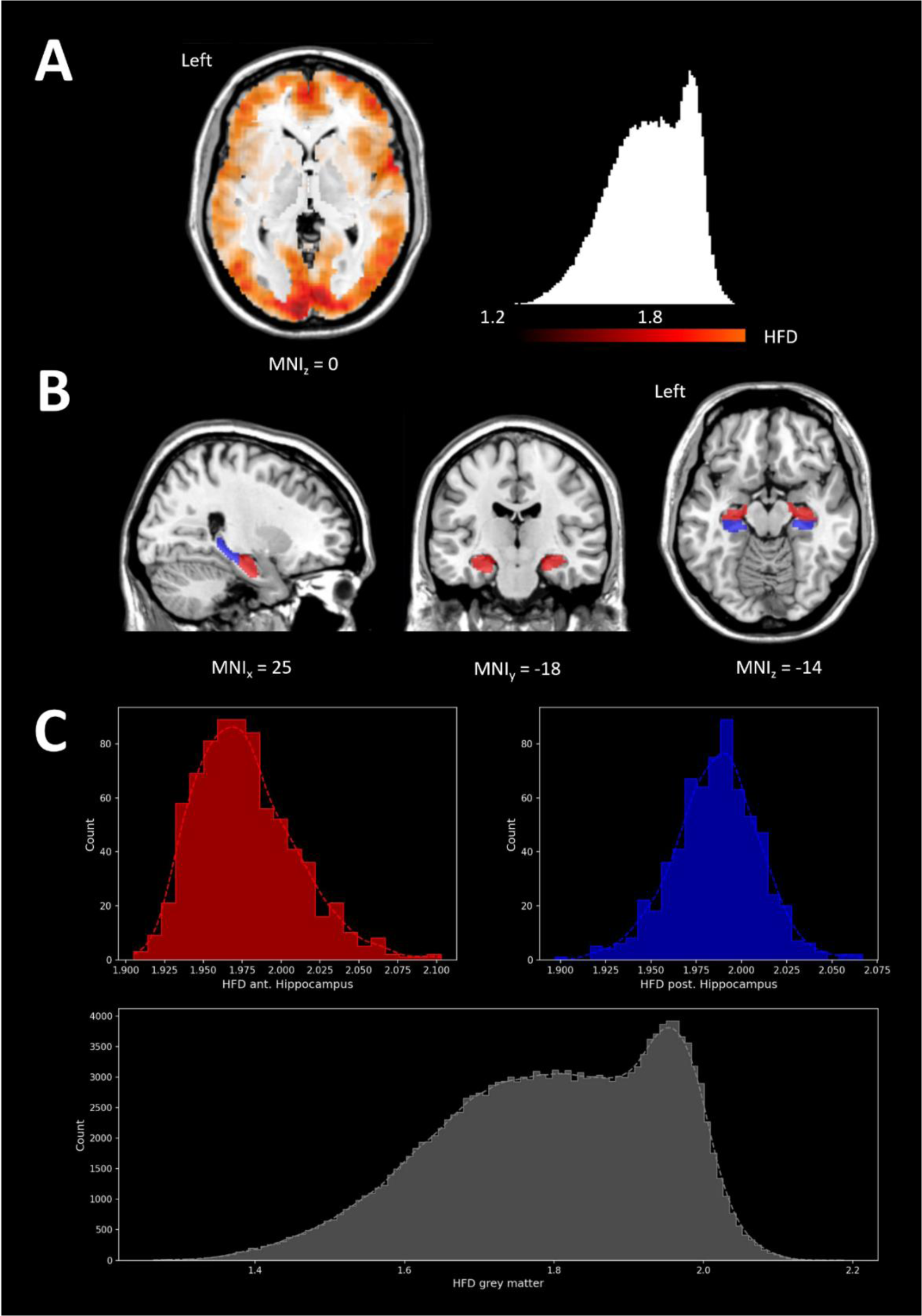
HFD distribution and visualization in a sample subject. **A:** Color-coded HFD map of grey matter. **B:** Binary masks of anterior (red) and posterior (blue) parts of the hippocampi. **C:** HFD Histogram of total grey matter and anterior/posterior part of the hippocampi. L: left; MNI: Montreal Neurological Institute space.

For computation of anterior and posterior hippocampi, we used the uncal apex as a standard for landmark-based segmentation of the bilateral hippocampi obtained by the Automatic Anatomical Labelling (AAL) atlas (Tzourio-Mazoyer et al., 2002) by defining a separation plane in standard MNI space of y = -21 (Bracht et al., 2023; Poppenk et al., 2013). We extracted mean values of the anterior and posterior bilateral hippocampi for HFD using Matlab.

### Calculation of fractal dimension using structural imaging

We performed structural MRI data pre-processing of MP2RAGE images using the Computational Anatomy Toolbox (CAT12, http://www.neuro.unijena.de/cat/). We used standard fully automated pipelines for processing surface-based morphometry and reconstruction of the surface (Dahnke et al., 2013; Gaser et al., 2022). For measurement of FD-CM, CAT12 implemented an approach using spherical harmonic reconstructions (Yotter et al., 2011a; Yotter et al., 2011b). The surface shape of the brain was reconstructed multiple times by increasing band-width of frequency of spherical harmonic reconstructions. Local FD-CM was calculated by finding the slope of a double logarithmic plot regressing area versus dimension with pairs {ln(surface area), ln (scale of measurement)}. Prior to the second-level analyses, local FD-CM information was re-parameterized into a standard coordinate system across all subjects and smoothed with a recommended large Gaussian full width at half maximum kernel of 20 mm. See figure 2.

**Figure 2:**
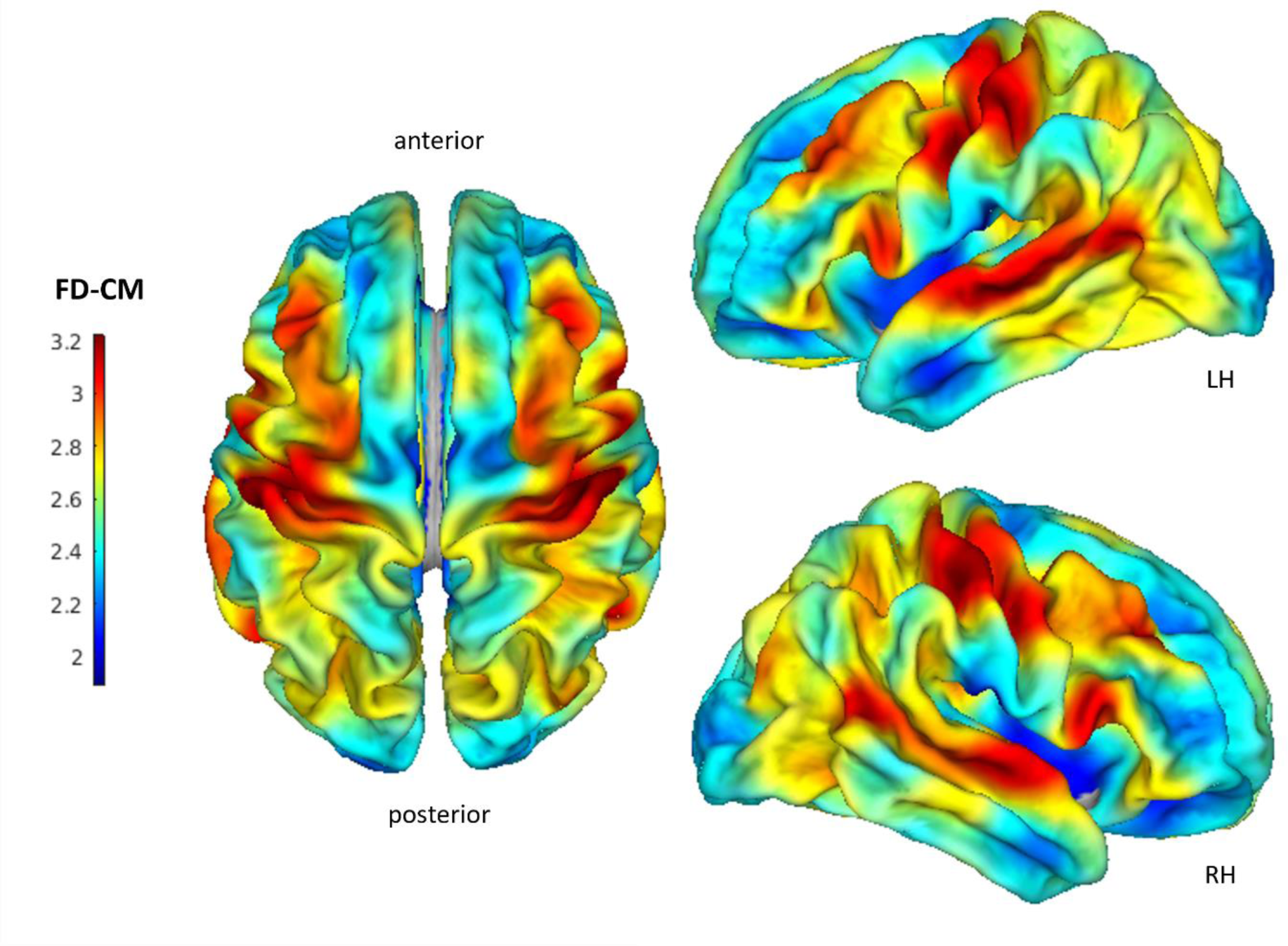
Overall mean of FD-CM. LH: left hemisphere; RH: right hemisphere.

## Statistical analyses

### Primary outcome measure

We used the Statistical Package for Social Sciences SPSS 29.0 (SPSS, Inc., Chicago, Illinois) to analyze HFD. We measured reliability of hippocampal HFD measurements within HC using Cronbach’s α and intraclass correlation coefficient (ICC) (Vaz et al., 2013). To investigate if time effects between the 2 MRI-scans differ between ECT-patients and HC, two separate repeated measures ANCOVAs with the independent variable group (ECT, HC), the within subject factors timepoint (baseline, follow-up) and hemisphere (left, right), the covariates age and sex and the dependent variables HFD (anterior and posterior hippocampus) were calculated. Significant group × time effects were followed up using post hoc paired t-tests comparing HFD.

### Secondary outcome measures

To investigate if there are associations between HFD changes over time and clinical improvement (relative changes between baseline (bl) and follow-up (fu): HAMD_%Δ_ = 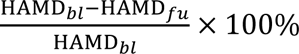.) within patients, we calculated exploratory Spearman correlations. To investigate the predictive value of baseline hippocampal HFD, we performed 4 separate regression analyses (anterior, posterior, left and right hippocampi), adjusting for age, sex and global HFD. P-values were Bonferroni corrected 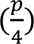.

To determine the predictive value of baseline FD-CM for HAMD reduction during the ECT-series a whole-brain multiple regression analysis was performed using baseline measures of FD-CM, adjusting age, sex and total intracranial volume, as covariates of no interest. Multiple regression was performed using the cortical parcellations of the Destrieux (aparc.a2009s) atlas (Destrieux et al., 2010). Inference statistics were done with a peak-level threshold of p < 0.05 and a Holm-Bonferroni correction of p < 0.05.

Additional exploratory analyses

In addition, we performed exploratory analyses regarding group differences at baseline, associations between the complexity measures HFD, FD-CM, and conventional structural and functional neuroimaging measures, and the predictive value of FD-CM on hippocampal HFD-changes (see supplementary material).

## Results

### Primary outcome measure

All hippocampal HFD values within HC showed no change over time with a good Test–Retest reliability as measured by Cronbachs’α and ICC (see table 2). Longitudinal analyses revealed significant group × time interactions for anterior hippocampal HFD (F_1,36_ = 5.071, p < 0.037, η^2^ =0.123) and posterior hippocampal HFD (F_1,36_ = 6.001, p < 0.019, η^2^ =0.143). Follow up paired t-tests revealed significant increase of HFD values over time in the left anterior and bilateral posterior hippocampus within ECT patients (see table 3 and figure 3).

**Figure 3:**
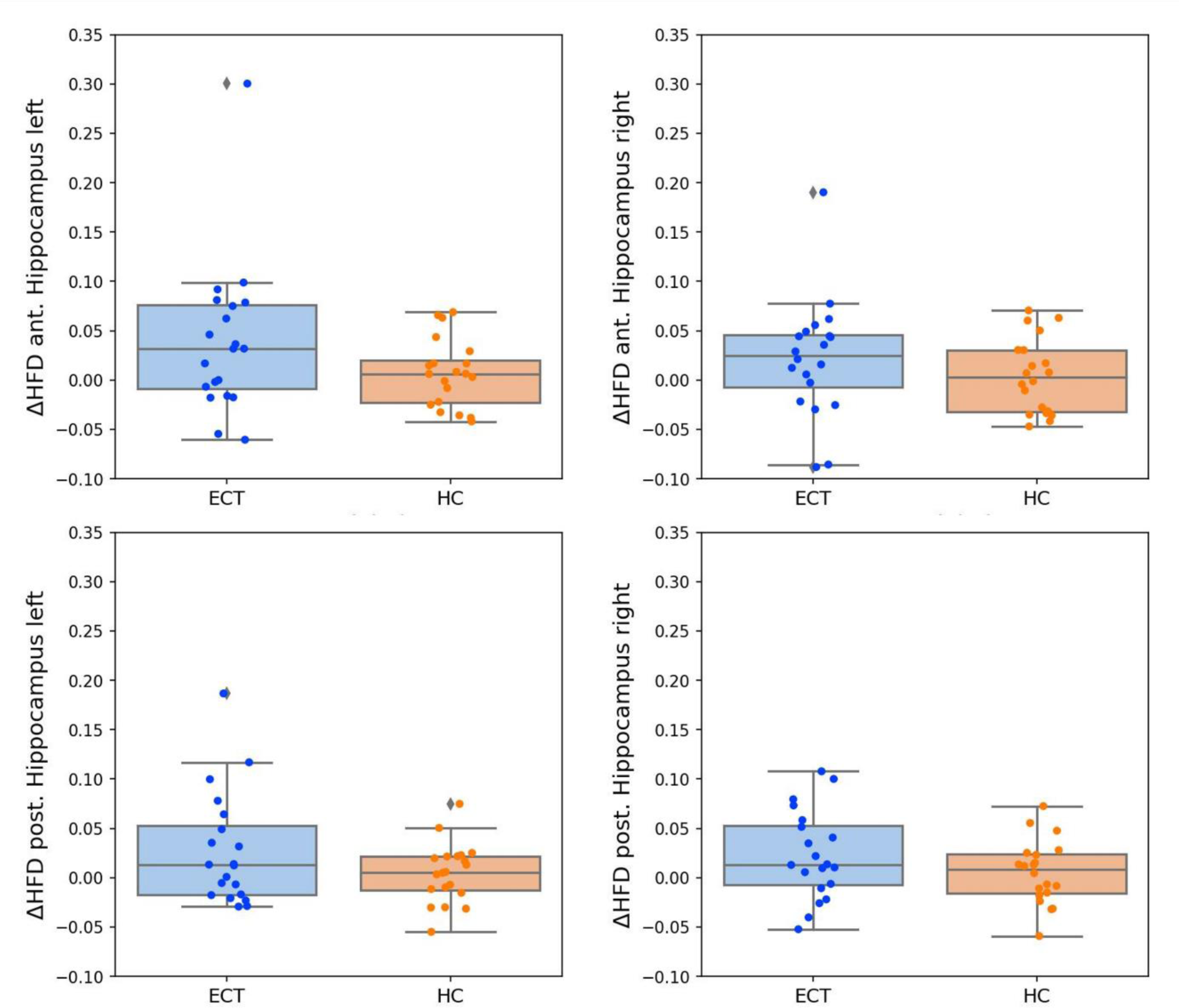
Hippocampal group differences in HFD.

**Table 2:** Test-Retest reliability for hippocampal HFD in healthy controls.

| HFD | T1 mean (SD) | T2 mean (SD) | Cronbach's $\alpha$ | ICC | \| $\Delta$ Mean \| | \| $\Delta$ SD \| |
| --- | --- | --- | --- | --- | --- | --- |
| Ant. hippocampus left | 1.928 (0.043) | 1.935 (0.05) | 0.844 | 0.732 | 0.0069 | 0.0344 |
| Ant. hippocampus right | 1.923 (0.046) | 1.927 (0.054) | 0.837 | 0.72 | 0.0041 | 0.0375 |
| Post. hippocampus left | 1.933 (0.037) | 1.937 (0.05) | 0.868 | 0.767 | 0.0045 | 0.03 |
| Post. hippocampus right | 1.927 (0.038) | 1.933 (0.045) | 0.825 | 0.702 | 0.0052 | 0.0321 |

HFD: Higuchi's fractal dimension; ICC: intraclass correlation coefficient; SD: standard deviation;
tion; T1: first MRI scan; T2: second MRI scan; $\Delta$ : difference.

**Table 3:** Post hoc paired t-tests for anterior and posterior hippocampal HFD in baseline vs. follow-up.

| Hemisphere | ECT | HC |
| --- | --- | --- |
| **HFD of anterior hippocampus** |  |  |
| **left** | $T_{19} = -2.240, p = 0.037$ | $T_{19} = -0.895, p = 0.554$ |
| **right** | $T_{19} = -1.617, p = 0.122$ | $T_{19} = -0.495, p = 0.627$ |
| **HFD of posterior hippocampus** |  |  |
| **left** | $T_{19} = -2.166, p = 0.043$ | $T_{19} = -0.667, p = 0.513$ |
| **right** | $T_{19} = -2.303, p = 0.033$ | $T_{19} = -0.724, p = 0.478$ |

### Secondary outcome measures

Within patients, clinical improvement in HAMD correlated positively with HFD increase in the left anterior hippocampus (r = 0.49, p = 0.03), but not in other hippocampal structures (see figure 4). Within ECT patients higher baseline FD-CM in the left temporal pole predicted clinical improvement in the ECT group as assessed with reduction in total HAMD scores. See table 4 and figure 5. Baseline hippocampal HFD values did not predict clinical improvement in HAMD after Bonferroni correction (see table 5).

**Figure 4:**
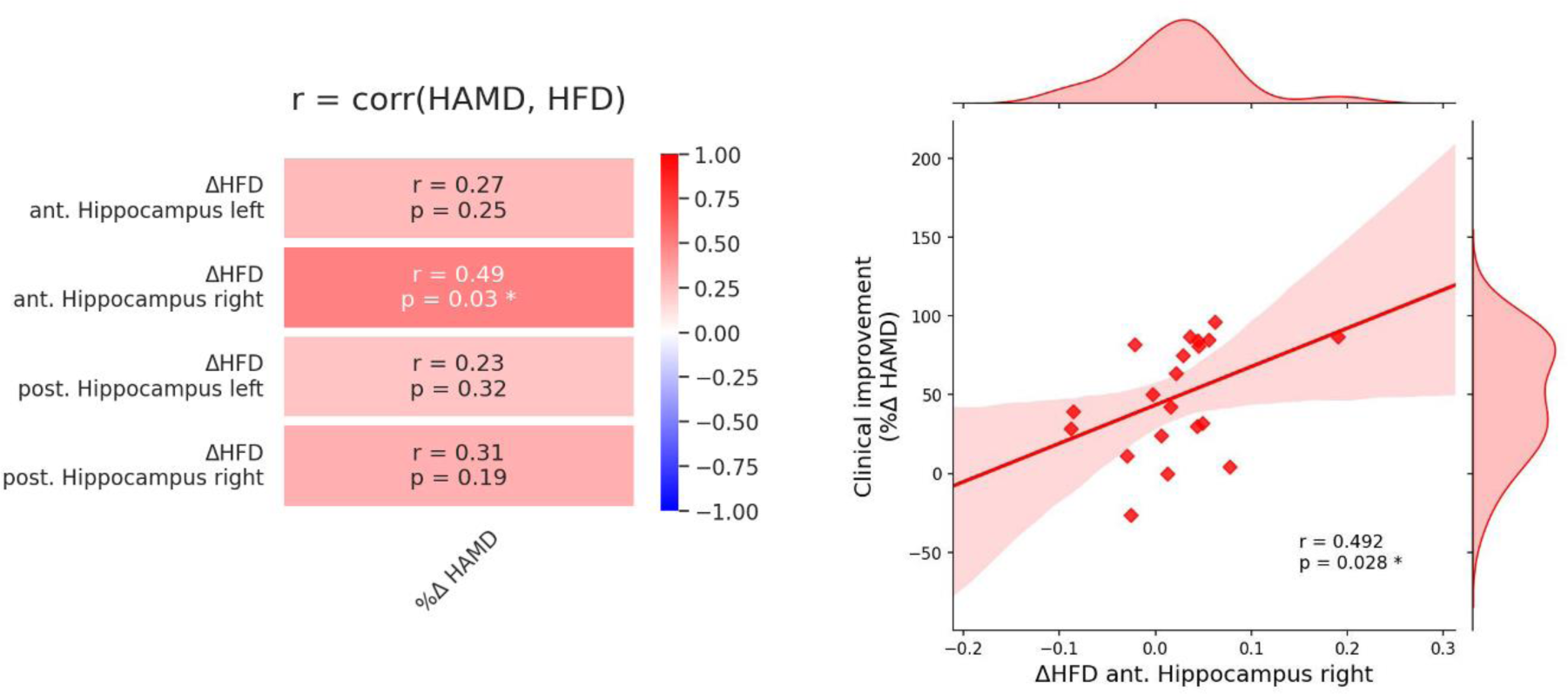
Positive association of clinical improvement and increase in HFD in the right anterior hippocampus.

**Figure 5:**
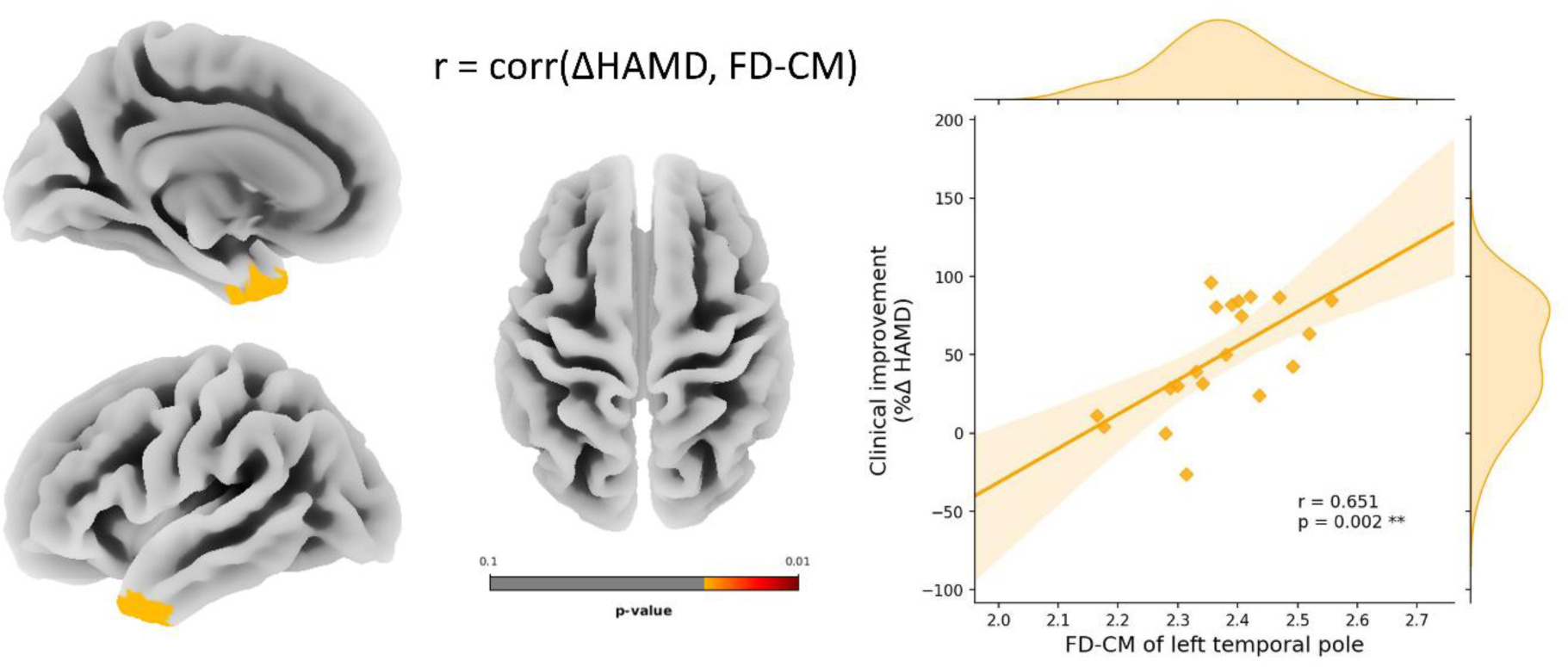
Positive association of the baseline FD-CM of the temporal pole and clinical improvement.

**Table 4:** Significant association of clinical improvement (%Δ HAMD) and baseline FD-CM.

| Direction | Region<br>(aparc_a2009s) | Hemisphere | T-value | Holm-Bonfer-<br>roni p-value |
| --- | --- | --- | --- | --- |
| positive | temporal pole | Left | 3.324 | 0.020 |
| negative | none |  |  |  |

aparc.a2009s: Destrieux atlas.

**Table 5:** Association of clinical improvement (%Δ HAMD) and baseline HFD.

| Baseline HFD | T-value | p-value | Bonferroni correction $\frac{p}{4}$ |
| --- | --- | --- | --- |
| Anterior hippocampus left | -1.805 | 0.091 | 0.364 |
| Anterior hippocampus right | -2.305 | 0.036* | 0.144 |
| Posterior hippocampus left | -2.351 | 0.033* | 0.132 |
| Posterior hippocampus right | -1.339 | 0.201 | 0.804 |

\*p < 0.05

## Discussion

Our study aimed to investigate neuroplasticity in patients with a current depressive episode undergoing ECT treatment by applying two complexity measurements of functional and structural MRI. This is the first study that investigates changes in HFD in a neuropsychiatric population using rs-fMRI. Through calculation of HFD, a functional metric of complexity, we demonstrated an ECT-induced increase in time-series complexity in bilateral hippocampi in patients. The increase in the right anterior hippocampus was positively associated with clinical improvement. In addition, by analyzing baseline FD-CM of structural data, we found that higher complexity of the left temporal pole was predictive for treatment response.

Our main finding was an ECT-induced increase in time-series complexity in bilateral anterior and left posterior hippocampi as assessed with HFD. Increase in the HFD in the right anterior hippocampus was associated with reductions in overall depression severity. Our results suggest a differential role of the anterior and the posterior hippocampi for treatment outcome and complement previous research separating these hippocampal compartments (Bracht et al., 2023; Leaver et al., 2019; Leaver et al., 2021). Our finding of an association between an increase of HFD in the right anterior hippocampus and clinical improvements fits the finding of (Leaver et al., 2021) who reported an ECT-induced volume increase in the right anterior hippocampus that was specific for ECT-treatment responders. However, Leaver et al. also reported that an increase in cerebral blood flow (CBF) in ECT-responders was located in the right middle and left posterior hippocampus (Leaver et al., 2021). Our previous analysis of this sample revealed an association between volume increase of the right posterior hippocampus and clinical improvement (Bracht et al., 2023). Thus, associations between volumetric and functional changes of hippocampal subcompartments and clinical response remain to be elucidated.

In addition to our finding of changes of HFD, we identified predictors of treatment outcome using FD-CM, a structural measure of complexity. Applying a multiple regression analysis of the whole brain, we found that higher FD-CM in the left temporal pole predicted clinical improvement following an ECT-index series. The identification of a structural measure located in the temporal pole makes sense because it is near the anterior hippocampus a region with connection pathways that have been linked to ECT-treatment response (Kubicki et al., 2019) and to structural grey matter remodeling in treatment responders (Leaver et al., 2021). In addition, volume reductions in the temporal pole have been reported in patients with MDD and BD (Neves et al., 2015; Webb et al., 2014). FD-CM is a measure of complexity that incorporates structural features such as gyrification, folding frequency, sulcal depth, convolution and gyral shape. Sulcal patterns are mainly determined before birth and stable across the lifespan (Cachia et al., 2016; Tissier et al., 2018), and are linked to cognition (Cachia et al., 2021). Thus, there may be strong impact on individual developmental processes related to genetics (Huang et al., 2023). If replicated in larger samples, FD-CM may contribute to distinguish ECT-treatment responders to non-responders in the future.

Our results provide further support for concepts that link hippocampal neuroplasticity to ECT treatment response. However, neuroimaging does not provide definite answers on the underlying neurobiological processes. Animal studies have demonstrated that seizures induced by electroconvulsive shocks are associated with enhanced hippocampal proliferation of neural stemlike cells, synapse formation, gliogenesis and angiogenesis (Chen et al., 2009; Hellsten et al., 2005; Newton et al., 2006; Olesen et al., 2017; Scott et al., 2000; Segi-Nishida et al., 2008; Wennström et al., 2004). The exact mechanism through which ECT enhances neurogenesis is not fully understood, but it likely involves the stimulation of various neurotrophic factors (e.g. brain-derived neurotrophic factor (BDNF), endothelial growth factor (VEGF) and basic fibroblast growth factor) which are believed to modulate hippocampal circuitries (Bolwig, 2011; Newton et al., 2003; Ueno et al., 2019). In humans, in contrast to the well-studied and replicated volumetric changes in the hippocampus, findings regarding neurotrophic factors are still limited. One recent ECT-study has shown, that hippocampal volume increase is particularly associated with VEGF (Van Den Bossche et al., 2019). Furthermore, ECT-induced hippocampal volume increase may be related to the dose of ECT-sessions and to electrode placement. However, recent research suggests that these factors may not only be related to antidepressive response but also to cognitive side effects (Argyelan et al., 2021; Bracht et al., 2023; Joshi et al., 2016; Leaver et al., 2022; Subramanian et al., 2022).

Overall, the search for further hippocampal and temporal lobe metrics continues to be of interest, as classical morphology and connectivity analyses to date have not yielded a conclusive picture (Gryglewski et al., 2021; Oltedal et al., 2018). In our data, we used HFD, a measure of functional complexity that so far has not been applied to ECT-research. It is worth mentioning that identified changes did not correlate with volumetric increases after the ECT-index series (see supplementary material S2, and figure S2). This suggests that HFD-assessed functional complexity is independent from MRI-assessed volumetric measures. Furthermore, it supports assumptions that the neurobiology of treatment response is too complex to be simply attributed to volumetric measures. Further research is needed to understand what the HFD measure represents in rs-fMRI. The functional aspects of the hippocampus in ECT treatment are far from being fully understood. It has been proposed, that neuroplastic changes in the hippocampus and its interactions with other brain regions, such as the thalamocortical and cerebellar networks, may play a role in the antidepressant response to ECT (Leaver et al., 2021). In a previous study, we proposed that remodeling of structural and functional connectivity between the hippocampus and the self-referential default mode network may be associated with a reduction in rumination, a core feature of depression (Denier et al., 2023). Overall, it is likely that hippocampal communication behaviour is becoming more complex, which may be due to neurogenesis, synaptogenesis and dendritogenesis (Sartorius et al., 2022). This in turn may be associated with clinical improvements in depression pathophysiology, which is likely related to hippocampal pathology (Bracht et al., 2022b; Schmaal et al., 2016).

While to the best of our knowledge HFD has not been applied in rs-fMRI in mood disorders, researchers applied the Hurst Exponent (HE), another mathematical metric of complexity. By comparing HFD and HE, HFD focuses on the local self-similarity and roughness of a time series, and HE is more concerned with the long-range statistical dependence and self-similarity of data. They are used for different purposes and provide different insights into the structure and behaviour of time series data (Krakovská and Krakovská, 2016). By assessing temporal dynamic of brain activity HE of the ventromedial prefrontal cortex was positively associated with rumination and mediated the association between rumination and depression (Gao et al., 2023). Using machine learning classification approaches the HE metric also helped to distinguish between major depression and healthy controls (Wei et al., 2013) and between remitted and current major depression (Jing et al., 2017). This highlights the potential of complexity measures for understanding the neurobiology of depression and remission plasticity.

Our study has certain limitations. Firstly, the sample size is modest and findings warrant replication in larger data sets (e.g. combining data with help of consortia (Oltedal et al., 2017)). Secondly, the population of depressed patients exhibits heterogeneity in terms of diagnoses, encompassing both MDD and BD, as well as variations in clinical characteristics such as the severity and duration of episode. However, this reflects clinical practice and therefore increases external validity. Thirdly, investigated measures of HFD and FD-CM are complex and do not allow for a straightforward neurobiological interpretation. Additional comprehensive research is imperative to deepen our comprehension of the interplay between HFD and FD-CM and conventional metrics such as cortical thickness or functional connectivity.

In conclusion, this study marks the pioneering exploration of HFD and FD-CM in the context of mood disorders and ECT. Our findings revealed a bilateral HFD increase in anterior and posterior hippocampi. Notably, the increase in HFD in the right anterior hippocampus was associated with clinical improvements. In addition, FD-CM measurements in the left temporal pole may be predictive for treatment response. Collectively, the utilization of complexity measurements such as HFD and FD-CM represent a novel and intriguing avenue for investigating neuropsychiatric disorders, particularly in patients undergoing ECT-treatment. Future studies with larger datasets are needed to further assess their predictive capabilities and associations with clinical improvements more comprehensively.

## Data Availability

All data produced in the present study are available upon reasonable request to the authors

## Acknowledgements

This study received funding from the Robert Enke Foundation and the Novartis Foundation for medical biological research (to Tobias Bracht and Sebastian Walther) and the Swiss Life Foundation (to Tobias Bracht). Niklaus Denier and Tobias Bracht were both funded independently by a grant from the Adrian et Simone Frutiger Foundation.

