## Supplementary Material for "Analyzing Fractal Dimension in Electroconvulsive Therapy: Unraveling Complexity in Structural and Functional Neuroimaging"

**Supplementary material S1: group comparisons**

Baseline group comparisons in HFD were performed with two separate mixed-model ANCOVAs with the independent variable group (ECT, HC), the within subject factor hemisphere (left, right), the covariates age and sex and the dependent variables HFD (anterior and posterior hippocampus) were calculated. Baseline analyses for HFD revealed no significant group effect for anterior hippocampal HFD ($F_{1,36} = 1.141$, $p = 0.293$) and posterior hippocampal HFD ($F_{1,36} = 2.180$, $p = 0.149$) (see supplementary figure S1).

To determine baseline group differences in FD-CM a two-sample t-test was performed using baseline FD-CM through the batch-mode implemented in CAT12, adjusting for total intracranial volume (TIV), age and sex as covariates of no interest. Inference statistics were done with a peak-level threshold of $p < 0.05$ and a Holm-Bonferroni correction of $p < 0.05$. No significant baseline FD-CM group differences survived Holm-Bonferroni correction in both directions.

**Figure S1:** Boxplot of hippocampal HFD measurements for baseline (T1) and follow-up (T2) assessment.

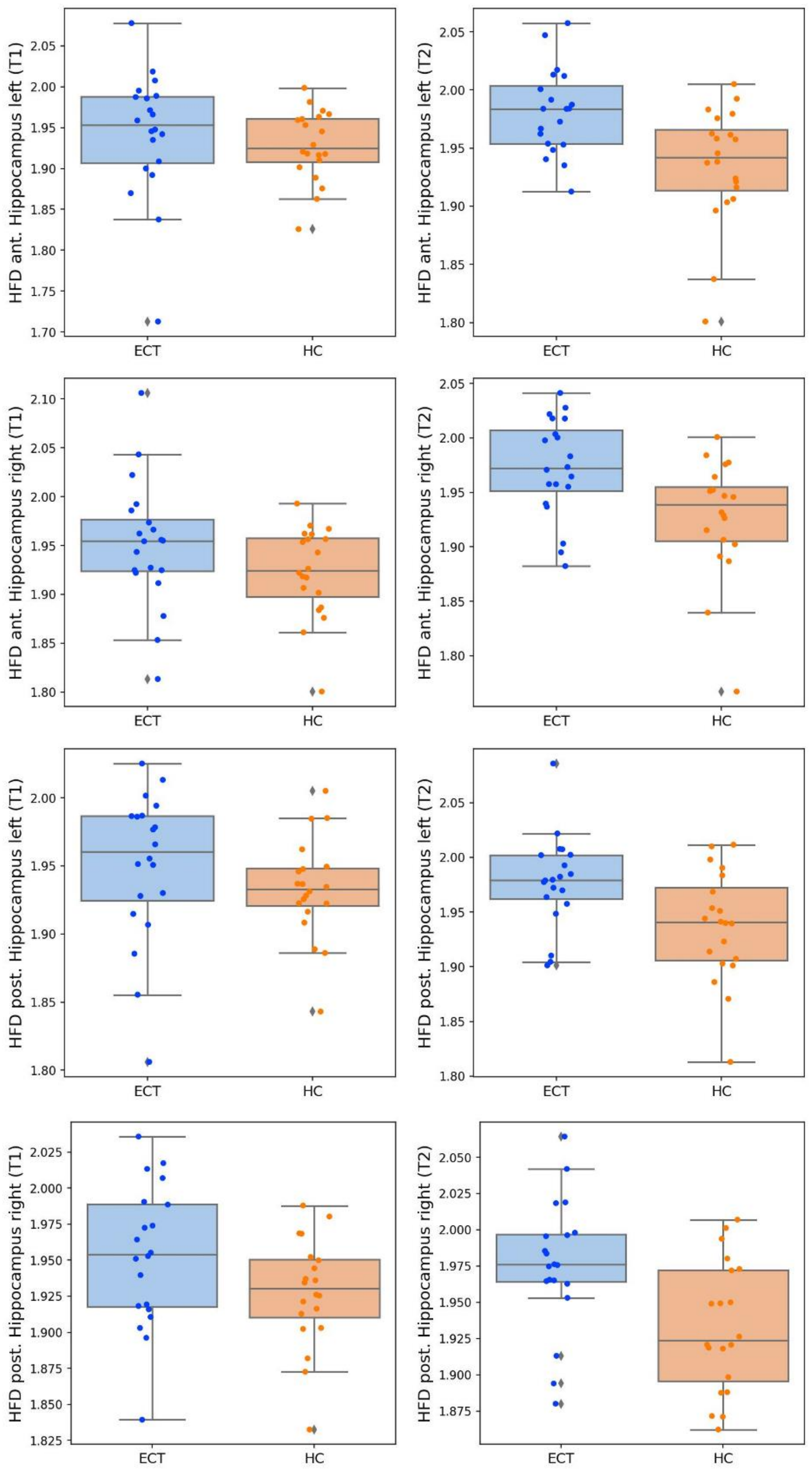

**Supplementary material S2: Associations between HFD, cerebral blood flow (CBF), and hippocampal volume**

To better understand HFD changes, we also performed Spearman correlations of already published volumetric and cerebral blood flow (CBF) findings (Bracht et al. 2023) with HFD changes over time. We found no significant correlations of already published volumetric and CBF increases after ECT-index series and HFD changes (see supplementary figures S2 and S3).

**Supplementary Figure S2:** Spearman correlations of hippocampal changes in HFD and volume (Bracht et al. 2023)

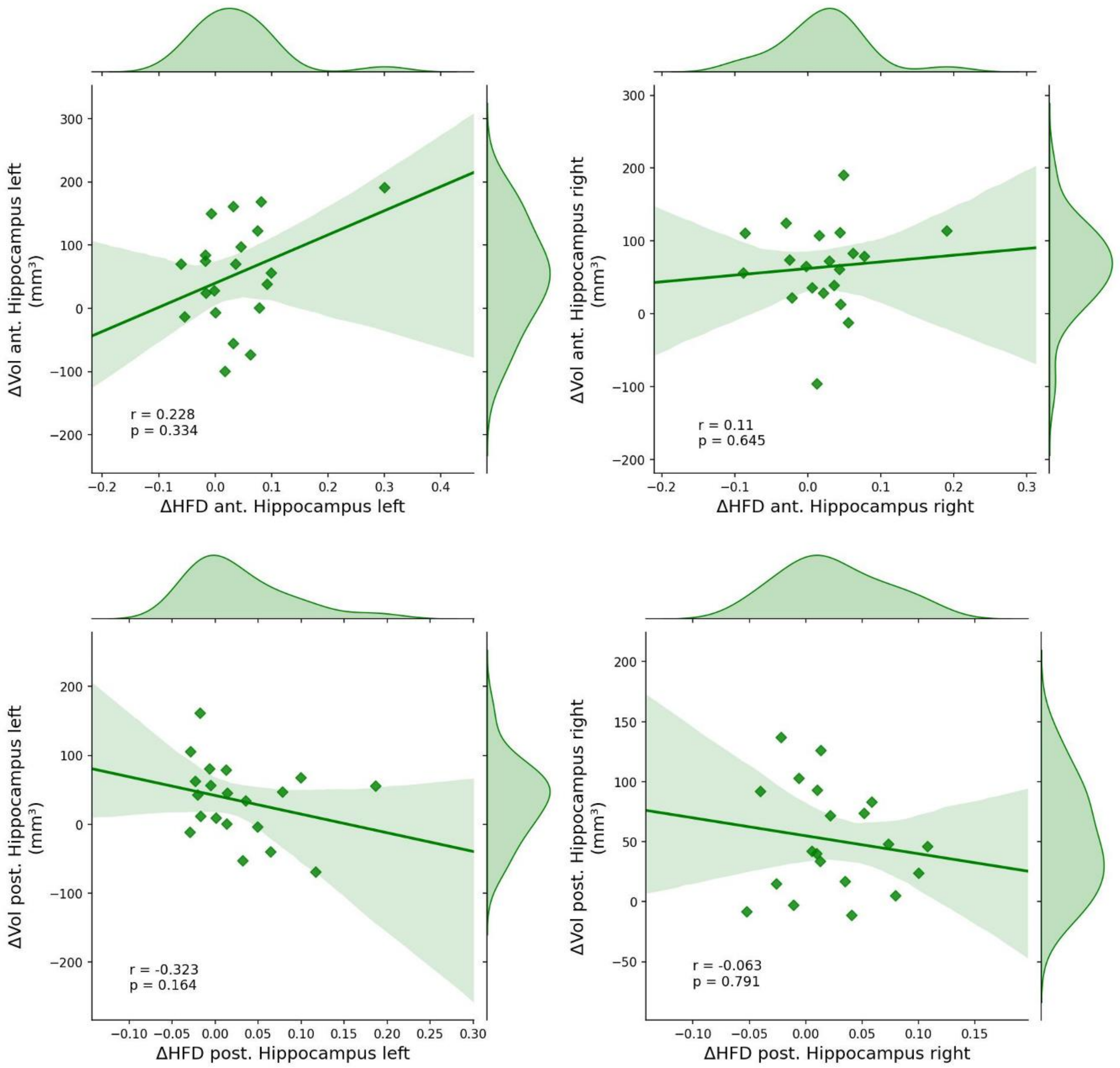

**Supplementary Figure S3:** Spearman correlations of hippocampal changes in HFD and CBF (Bracht et al. 2023)

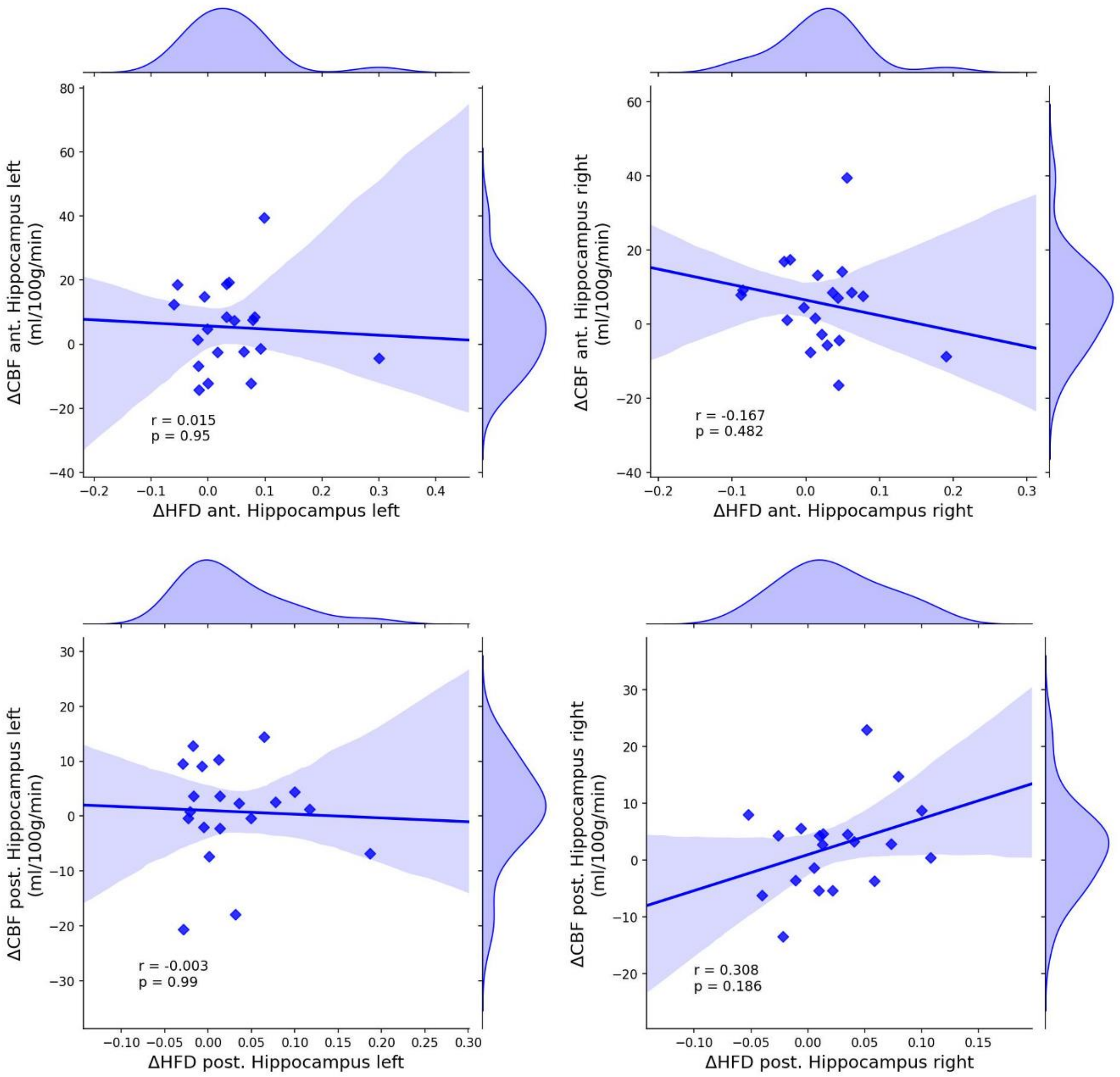

**Supplementary material S3: Investigation of additional resting-state fMRI metrics**

We computed in addition to HFD two rs-fMRI metrics including intrinsic connectivity (IC) and fractional amplitude of low-frequency fluctuations (fALFF). IC is defined as the mean square of correlation coefficients ($r$) between a voxel ($x$) and all other voxels ($y$) within the brain ($B$): $\mathrm{IC}(x) = \left(\int_y^B r^2(x,y)dy\right)^{1/2}$. In contrast to degree-based network metrics, the IC power approach considers not only the number of connections but also its connectivity strength (Martuzzi et al. 2011). The fALFF metric was calculated by transforming the fMRI time series in each voxel using the Fourier transform function to the frequency domain and calculating the power spectrum. The ratio of the power of the low frequency fluctuations (0.01-0.10 Hz) was calculated relative to the full frequency range (Zou et al. 2008). We extracted mean values of the anterior and posterior bilateral hippocampi for IC and fALFF using Matlab.

We used the Statistical Package for Social Sciences SPSS 29.0 (SPSS, Inc., Chicago, Illinois) and paired t-tests to analyze HFD changes over time within groups.

We also found no significant longitudinal differences for IC and fALFF for HC and ECT patients (see supplementary table S1).

**Supplementary Table S1:** Post hoc paired t-tests for anterior and posterior hippocampal IC and fALFF in baseline vs. follow-up.

| **Hemisphere** | **ECT** | **HC** |
|---|---|---|
| **IC of anterior hippocampus** | | |
| Left | $T_{19} = 0.038$, $p = 0.970$ | $T_{19} = 1.713$, $p = 0.103$ |
| Right | $T_{19} = 0.175$, $p = 0.863$ | $T_{19} = 0.426$, $p = 0.675$ |
| **IC of posterior hippocampus** | | |
| Left | $T_{19} = 0.077$, $p = 0.939$ | $T_{19} = 0.868$, $p = 0.396$ |
| Right | $T_{19} = -0.022$, $p = 0.983$ | $T_{19} = 0.940$, $p = 0.359$ |
| **fALFF of anterior hippocampus** | | |
| Left | $T_{19} = -0.807$, $p = 0.430$ | $T_{19} = 0.468$, $p = 0.645$ |
| Right | $T_{19} = -0.104$, $p = 0.918$ | $T_{19} = -0.950$, $p = 0.354$ |
| **fALFF of posterior hippocampus** | | |
| Left | $T_{19} = -1.040$, $p = 0.311$ | $T_{19} = -0.731$, $p = 0.473$ |
| right | $T_{19} = 1.102$, $p = 0.284$ | $T_{19} = 0.680$, $p = 0.505$ |

fALFF: fractional amplitude of low IC: intrinsic connectivity.

**Supplementary Material S4: Investigation of predictive value of FD-CM on hippocampal HFD changes.**

We investigated possible predictive value of FD-CM baseline values for hippocampal HFD changes over time with Spearman correlations. Baseline FD-CM of the left temporal pole was not associated with hippocampal HFD changes of the left hemisphere (see supplementary figure S4).

**Supplementary figure S4: Investigation of predictive value of FD-CM and hippocampal HFD changes.**

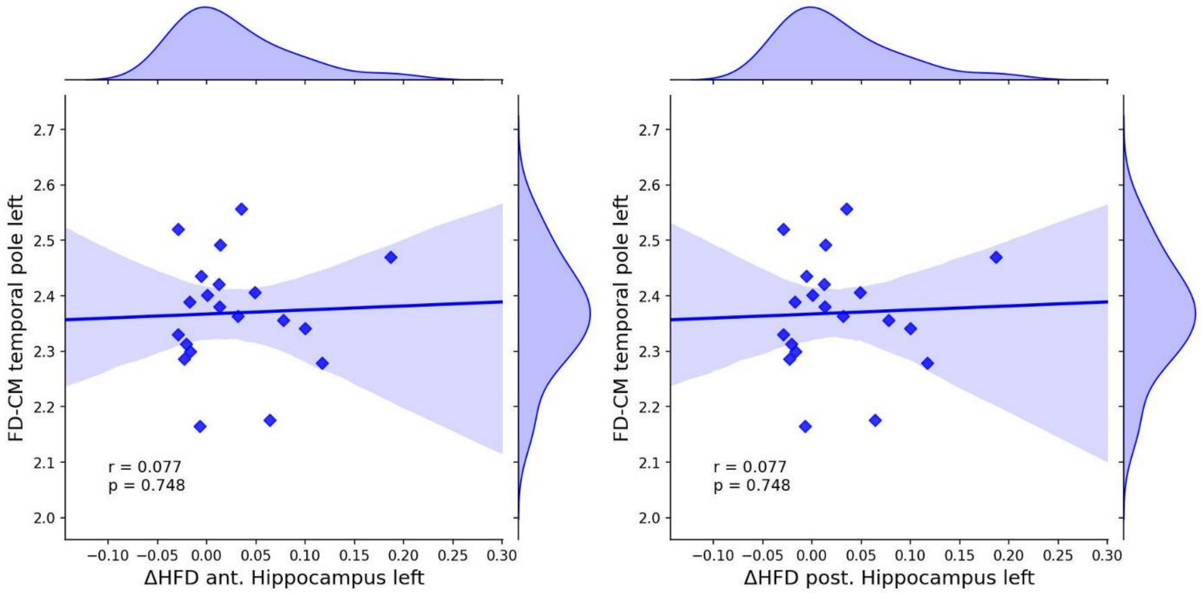
